# Risk of Atherosclerotic Cardiovascular Disease Hospitalizations after COPD Hospitalization among Older Adults

**DOI:** 10.1101/2023.12.19.23300254

**Authors:** Christopher L. Mosher, Oyomoare L. Osazuwa-Peters, Michael G. Nanna, Neil R. MacIntyre, Loretta G. Que, W. Schuyler Jones, Scott M. Palmer, Emily C. O’Brien

## Abstract

**BACKGROUND:** Meta-analyses have suggested the risk of atherosclerotic cardiovascular disease (ASCVD) events is significantly higher after a chronic obstructive pulmonary disease (COPD) exacerbation. However, these studies have been limited to highly selected patient populations potentially not generalizable to the broader population of COPD.

**METHODS:** We assessed the risk of ASCVD hospitalizations after COPD hospitalization compared to before COPD hospitalization and identified patient factors associated with ASCVD hospitalizations after COPD hospitalization. This retrospective cohort study used claims data from 920,550 Medicare beneficiaries hospitalized for COPD from 2016-2019 in the US. The primary outcome was risk of a ASCVD hospitalization composite outcome (myocardial infarction, percutaneous coronary intervention, coronary artery by-pass graft surgery, stroke, or transient ischemic attack) in the 1 year after-COPD hospitalization relative to the 1 year before-COPD hospitalization. Time from discharge to a composite ASCVD hospitalization outcome was modeled using an extension of the Cox Proportional-Hazards model, the Anderson-Gill model with adjustment for patient characteristics. Additional analyses evaluated for interactions in subgroups and risk factors associated with the composite ASCVD hospitalization outcome.

**RESULTS:** Among 920,550 patients (mean age, 73 years) the hazard ratio estimate (HR; 95% CI) for the composite ASCVD hospitalization outcome after-COPD hospitalization vs before-COPD hospitalization was 0.99 (0.97, 1.02; p = 0.53) following adjustment. We observed 3 subgroups that were significantly associated with higher risk for ASCVD hospitalizations after COPD hospitalization: 76+ years old, women, COPD hospitalization severity. Among the 19 characteristics evaluated, 10 were significantly associated with higher risk of CVD events 1 year after COPD hospitalization with hyperlipidemia (2.78; 2.67, 2.90) and history of cardiovascular disease (1.77; 1.72 1.83) associated with the greatest risk.

**CONCLUSION:** Among Medicare beneficiaries hospitalized for COPD, the risk of ASCVD hospitalizations was not significantly increased after COPD-hospitalization relative to before-COPD hospitalization. Although, we identified age 76+ years old, female sex, and COPD hospitalization severity as high risk subgroups and 10 risk factors associated with increased risk of ASCVD events after-COPD hospitalization. Further research is needed to characterize the COPD exacerbation populations at highest ASCVD hospitalization risk.

## INTRODUCTION

Among the 16 million people living with chronic obstructive pulmonary disease (COPD) in the US^1^, over 700,000 hospitalizations for COPD exacerbations occurred annually between 2001 and 2012^2^. While COPD hospitalizations are already recognized as sentinel events associated with increased mortality^3^, increasing evidence suggests that patients who experience COPD exacerbations are at 3-to 10-fold increased risk of atherosclerotic cardiovascular disease (ASCVD) events following COPD exacerbations ^4–6^. The risk of ASCVD events appears greatest among those with a severe exacerbation requiring hospitalization and remains elevated at 12 months^6^. While there is growing evidence that the period directly following COPD exacerbations is critical for reducing ASCVD event risk, there is a paucity of data to guide development of interventions targeted to patients at highest risk during this period.

The current COPD post-hospitalization treatment paradigm focuses exclusively on pulmonary-specific strategies such as triple inhaler therapy, anti-inflammatory medications, and pulmonary rehabilitation^7^. Current guidelines acknowledge that patients already at high-risk for ischemic heart disease are at even greater risk of ASCVD events after COPD exacerbation^7^. However, there is currently no accepted guidance for mitigating this risk through evidence-based strategies, which may lead to potentially inappropriate undertreatment of high-risk patients following COPD exacerbations. In order to identify the populations most likely to benefit from intensified preventive strategies, the highest risk clinical phenotypes for future ASCVD hospitalizations after-COPD hospitalization must be characterized. Existing studies have been limited by restriction to patients with moderate obstructive lung disease^6^, use of non-adjudicated primary outcome data^4^, and studied in populations enriched for CVD^5, 6, 8^. Therefore, there is an urgent need to quantify the risk of ASCVD hospitalizations after COPD hospitalizations and identify ASCVD risk factors in a real-world population to inform the design of future interventional studies.

Our study focused on COPD and ASCVD hospitalizations as hospitalized patients seem to be at greatest risk of ASCVD events and high risk of future hospitalizations^6, 9, 10^. Our objectives were to 1) quantify the risk of ASCVD hospitalizations following COPD hospitalization compared to pre-COPD hospitalization baseline period, and 2) identify the patient factors associated with ASCVD hospitalizations after COPD hospitalization. We utilized Medicare claims data to quantify the risk of an ASCVD hospitalization composite outcome after COPD hospitalization and identify risk factors associated with ASCVD hospitalizations in a broad, generalizable population of older adults in the United States.

## METHODS

We performed a retrospective cohort study investigating the risk of atherosclerotic cardiovascular disease hospitalizations among older adults with COPD who were recently hospitalized for COPD. We utilized inpatient facility claims and corresponding Master Beneficiary Summary Files (Limited Data Set) for Medicare Part A fee-for-service beneficiaries from 2014 through 2019. The study was approved by the institutional review board (Pro00111343) at Duke Health, Durham, NC, USA.

Study cohort selection followed a similar approach as previously published in high-impact COPD literature using Medicare claims data^11, 12^ (supplementary Table S1). All patients met the following criteria: 1) hospitalized for a principal ICD-10 diagnosis of COPD or acute respiratory failure with secondary diagnosis of COPD exacerbation between January 1, 2016 and December 31, 2019; 2) ≥ 67 years old at the time of first hospitalization; 3) continuous enrollment in Medicare fee-for-service for 2 years prior to the index COPD hospitalization; 4) located in the continental USA (Figure 1). If multiple qualifying hospitalizations occurred, we selected the first qualifying COPD hospitalization.

**Figure 1:**
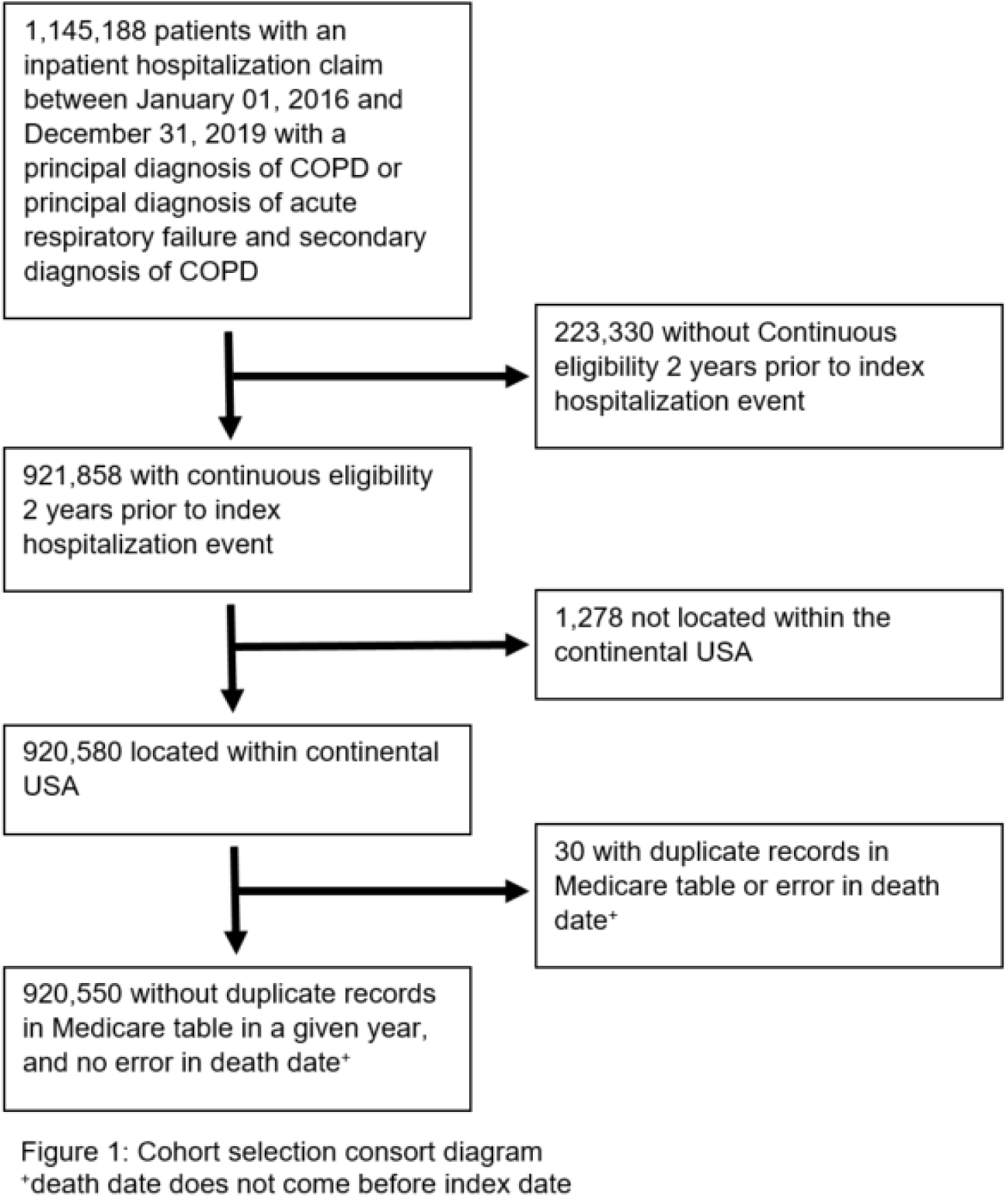
Consort study diagram

Baseline characteristics of patients at the time of first COPD hospitalization were described using frequencies and percentages for categorical variables and means (SD) and medians (first quartile (Q1), third quartile (Q3)) for continuous variables. The primary outcome was an ASCVD hospitalization composite outcome, defined by an inpatient claim with a primary diagnosis (i.e., DX1 field in claims data) of myocardial infarction (MI), percutaneous coronary intervention (PCI), coronary artery by-pass graft surgery (CABG), stroke, or transient ischemic attack (TIA) identified using previously validated algorithms (supplementary Table S2 for ICD-10 code algorithms). All ASCVD hospitalizations were included from index date through 1-year follow up after first COPD hospitalization except if MI, stroke, or TIA occurred on the same day as the day of admission for the first COPD hospitalization because they would not occupy the primary diagnosis field (DX1). PCI and CABG were included beginning on the day of admission for the first COPD hospitalization. MI, stroke, and TIA were included following the day of admission through the 1-year follow up period.

To mitigate confounding by patient characteristics, we compared the risk of ASCVD hospitalization in the 1-year following COPD hospitalization compared to the 1-year prior to COPD hospitalization (“baseline year”). We determined the index date as the date exactly one year before the date of admission for the first COPD hospitalization. All patients hospitalized for COPD were included, even if they did not survive to hospital discharge. Baseline covariates were determined in the 1-year prior to this index date using diagnostic codes from inpatient claims. Follow-up extended from the index date to the first of disenrollment, death, two years from index date, or the study end date (December 31, 2019) (Figure 2).

**Figure 2:**
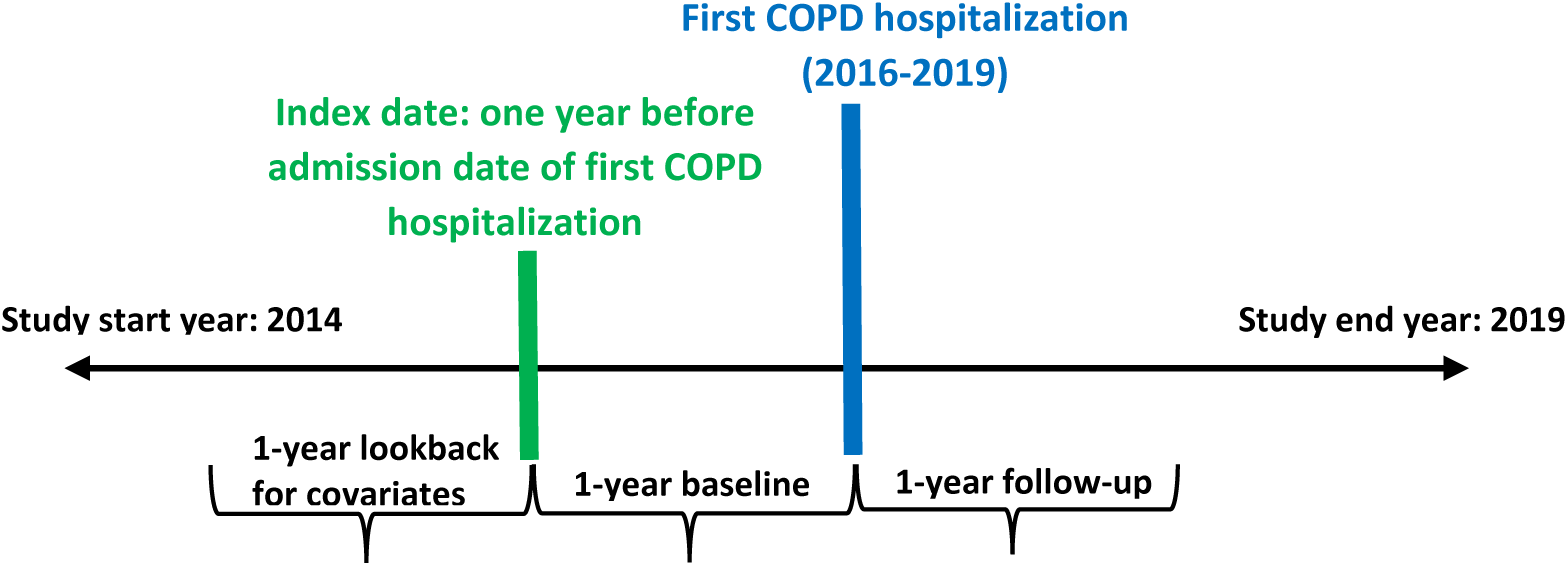
study schema

We calculated the cumulative incidence of the composite ASCVD hospitalization outcome in the year before and the year after COPD hospitalization using the cumulative incidence function method, which has the advantage of accounting for death as a competing risk^13^. The main study variable of interest was COPD hospitalization period, a time-varying variable that was 0 in the period before (i.e., pre-) and 1 in the period after (i.e., post) the index COPD hospitalization. We estimated the hazard ratio for post-COPD hospitalization vs. pre-COPD hospitalization period on repeated time-to-composite ASCVD hospitalizations over the entire two-year observation period using an extension of the Cox Proportional-Hazards model, the Anderson–Gill model^14^. In contrast to traditional Cox models that only model time to the first event, extended Cox models make use of all available data including but not limited to recurrent events, while accounting for the correlation between multiple events within subjects^14^. Extended Cox models have been shown to be more powerful to detect effects for repeated time-to-event health conditions^14^ such as the ASCVD hospitalization composite outcome.

Using Anderson-Gill models, we estimated HRs, 95% CIs and corresponding p-values for unadjusted models and models adjusting for age, sex, race, Medicaid dual eligibility (a surrogate maker for low socioeconomic status), comorbidities defined using claims^15, 16^ in the baseline period (hypertension, diabetes, hyperlipidemia, congestive heart failure, history of cardiovascular disease, cardiac arrhythmia, peripheral vascular disease, renal disease, valvular heart disease, dementia, metastatic cancer), index year, and frailty^17, 18^ (Table S3, S4). P-values < 0.05 were considered statistically significant in all analyses. We assessed the proportional hazard assumptions for all regression models when relevant using the Schoenfeld residual test (Table S5).

We tested for interactions between each of the following subgroups in separate Anderson-Gill models: prior diagnosis of CVD vs no prior CVD diagnosis, age 67-75 vs ≥76, frail^17, 18^ vs non-frail (Table S4), and men vs women. If a significant interaction was found, separate Anderson-Gill models within each subgroup were performed to estimate subgroup hazard ratios for the COPD hospitalization period variable.

We also examined whether the association between risk of ASCVD hospitalization and COPD hospitalization period differed depending on severity of the COPD hospitalization. In our dataset, we were unable to distinguish between non-invasive ventilation used for routine, nocturnal use (e.g., continuous positive airway pressure (CPAP)) and acute treatment for respiratory failure (e.g., bi-level positive airway pressure (BPAP)). Therefore, a COPD hospitalization that required treatment with invasive mechanical ventilation (IMV) was defined as a very severe COPD hospitalization (Table S6). Since COPD hospitalization occurred one year after index date (i.e., by the end of baseline period), we defined a 3-level time-varying categorical COPD hospitalization burden variable to capture this variation, such that COPD hospitalization burden could be defined as absent (i.e., did not occur during baseline period prior to hospitalization), severe defined as hospitalized but without treatment with IMV, and very severe defined as hospitalized with IMV treatment during index COPD hospitalization. Using Anderson-Gill models, we estimated adjusted hazard ratios for the 3-level categorical COPD burden variable, with pairwise comparisons among the groups (i.e., after-COPD hospitalization severe group vs. before-COPD hospitalization, after-COPD hospitalization very severe group vs. before-COPD hospitalization, and after-COPD hospitalization very severe group vs. after-COPD hospitalization severe group).

Further, we investigated risk factors associated with the ASCVD hospitalization composite outcome and evaluated if the associations changed over time by performing separate Cox proportional hazards regression models for time periods: 0-30 days, 0-90 days, 0-180 days, and 0-365 days following COPD hospitalization discharge date. For each time period, crude and adjusted associations were estimated between time-to-ASCVD hospitalization and potential risk factors.

## RESULTS

Among the 920,550 patients in the analysis cohort, the median (25^th^, 75^th^) age was 73 years old (67, 80), predominantly female (57%), white (87%), and 28% had dual Medicare and Medicaid eligibility. The most common comorbidities were hyperlipidemia (68%) and hypertension (32%). Nearly 10% of the population met criteria for very severe COPD hospitalization defined as requiring treatment with IMV. The median (25^th^, 75^th^) length of stay was 4 days (2, 6) (Table 1).

**Table 1:** Baseline characteristics of COPD cohort from 2016-2019.

|  |  |
| --- | --- |
| N | 920,550 |
| Age at index, years |  |
| Median (25th, 75th) | 73.0 (67.0, 80.0) |
| Mean (SD)) | 72.8 (10.6) |
| Age group at index, % |  |
| <65 | 170,176 (18.5) |
| 65-69 | 159,562 (17.3) |
| 70-74 | 179,458 (19.5) |
| 75-79 | 160,513 (17.4) |
| 80-84 | 125,241 (13.6) |
| 85+ | 125,600 (13.6) |
| Female, n (%) | 522,501 (56.8) |
| Race, n (%) |  |
| White | 801,841 (87.1) |
| Black | 82,925 (9.0) |
| Other/unknown | 35,784 (3.9) |
| US Census Region, n (%) |  |
| South | 405,432 (44.0) |
| Northeast | 234,642 (25.5) |
| Midwest | 124,683 (13.5) |
| West | 155,793 (16.9) |
| Dual eligibility | 263,848 (28.7) |
| Frailty, n (%) | 59,521 (6.5) |
| Comorbidities, n (%) |  |
| Hypertension | 297,527 (32.3) |
| Diabetes Mellitus | 140,419 (15.3) |
| Congestive Heart Failure | 137,387 (14.9) |
| History of Cardiovascular Disease | 168,531 (18.3) |
| Coronary Artery Disease | 161,857 (17.6) |
| History of Stroke | 26,891 (2.9) |
| Cardiac Arrhythmias | 146,342 (15.9) |
| Peripheral Vascular Disease | 62,602 (6.8) |
| Renal Disease | 98,811 (10.7) |
| Valvular Heart Disease | 48,800 (5.3) |
| Depression | 97,420 (10.6) |
| Hyperlipidemia | 626,534 (68.1) |
| Dementia | 24,566 (2.7) |
| Metastatic Solid Tumor | 4,785 (0.5) |
| Characteristics of index COPD hospitalization |  |
| Year of index COPD event, n (%) |  |
| 2016 | 306,909 (33.3) |
| 2017 | 276,617 (30.0) |
| 2018 | 182,970 (19.9) |
| 2019 | 154,054 (16.7) |
| Severity, n (%) | 84,704 (9.2) |
| Length of stay |  |
| Median (25th, 75th) | 4.0 (2.0, 6.0) |
| Mean (SD) | 5.4 (7.2) |
| Discharge location: home, n (%) | 636,842 (69.2) |

### Atherosclerotic cardiovascular disease hospitalizations and risk after COPD hospitalization

The total number of new cases of composite ASCVD hospitalizations (cumulative incidence %) in the 2-year follow-up period was 54,617 (6%). The most common hospitalizations were PCI 23,497 (2.6%) and stroke 18,121 (2%) (Table S7). The hazard ratio estimate for the ASCVD hospitalization composite outcome in the after-COPD hospitalization vs the before-COPD hospitalization was 1.06 (1.04, 1.08; p < 0.0001) without adjustment and 0.99 (0.97, 1.02; p = 0.53) following adjustment (Table 2).

**Table 2:** Crude and adjusted hazard ratio estimates from recurrent cox model.

| Variable | Crude Hazard Ratio (95% CI) | P | Adjusted* Hazard Ratio (95% CI) | P |
| --- | --- | --- | --- | --- |
| COPD hospitalization period |  |  |  |  |
| Before | Reference |  | Reference |  |
| After | 1.06 (1.04, 1.08) | <.0001 | 0.99 (0.97,1.02) | .53 |

\*Adjusted for age, sex, race, Medicaid dual eligibility, comorbidities (hypertension, diabetes, CHF, history of cardiovascular disease, cardiac arrhythmia, PVD, renal disease, VHD, depression, dementia, metastatic cancer, and hyperlipidemia), index year, and frailty

### Subgroups at risk for atherosclerotic cardiovascular disease hospitalizations after COPD hospitalization

We observed significant interactions between COPD hospitalization period, and age, sex, and COPD hospitalizations defined as very severe with hospitalizations for the ASCVD composite outcome pre– and post-COPD hospitalization (Table S8). In women, there was a 9% higher risk of the ASCVD composite outcome in after-COPD hospitalization relative to before. Conversely, in men, we observed a 9% lower risk of ASCVD hospitalizations post– vs. pre-COPD hospitalization. We observed interaction by age, with a 7% lower risk of ASCVD hospitalizations in age strata 67-75 years and a 6% higher risk in age strata 76+ in post-COPD hospitalization vs pre-COPD hospitalization (Figure 3). We found a 44% higher risk of ASCVD hospitalization comparing post-COPD very severe hospitalization (i.e., required use of invasive mechanical ventilation) vs. pre-COPD hospitalization and 50% higher risk of ASCVD hospitalization comparing post-COPD very severe hospitalization vs. post-COPD severe hospitalization (i.e., did not require invasive mechanical ventilation) (Figure 3).

**Figure 3:**
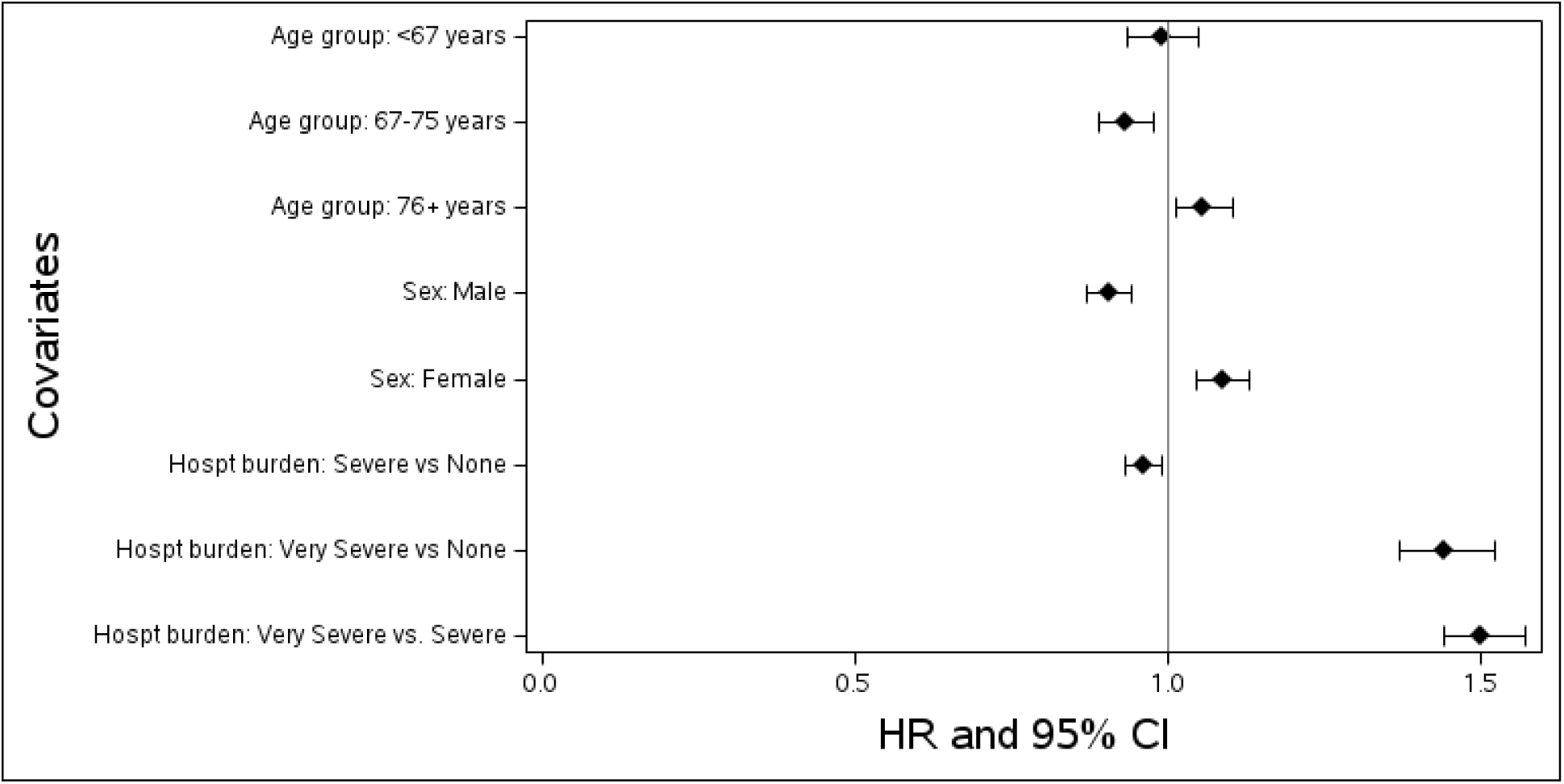
Forest plot showing subgroup specific adjusted hazard ratios for association of COPD time period (after vs before) and composite CVD outcome using a recurrent Cox regression model for subgroups that had significant interactions with COPD time period. Also showing adjusted hazard ratios for association of COPD hospitalization burden (i.e., hospt in plot; combination of COPD period and hospitalization severity) and composite CVD outcome using a recurrent Cox regression model. Note that there was no hospitalization burden in the before period, while in the after period, hospitalization burden could either be Severe or Very Severe. Each stratified model was adjusted as applicable for age, sex, race, Medicaid dual eligibility, comorbidities (hypertension, diabetes, CHF, history of cardiovascular disease, cardiac arrhythmia, PVD, renal disease, VHD, depression, dementia, metastatic cancer, and hyperlipidemia), index year, and frailty.

### Risk factors for atherosclerotic cardiovascular disease hospitalizations after COPD hospitalization

We identified risk factors for an ASCVD hospitalization composite outcome during the period following COPD hospitalization and evaluated changes in these associations over time. Among the 19 characteristics evaluated, 15 were significantly associated with risk of ASCVD hospitalizations after COPD hospitalization; 10 were associated with higher risk and 5 were associated with lower risk (Figure 4). For the majority of risk factors, the estimated risk of ASCVD hospitalizations remained consistent over time. However, the risk of ASCVD hospitalizations changed over time in history of cardiovascular disease, very severe COPD hospitalization, and hyperlipidemia. Compared to no CVD history, those with a history of CVD had higher risk of the ASCVD hospitalization composite outcome (HR; 95% CI) in the first 30 days following COPD hospitalization (2.21; 2.07,2.36) and despite a steady decline, remained elevated after 1 year (1.77; 1.72,1.83). Similarly, for those with very severe COPD hospitalization, the risk of ASCVD events was elevated in the first 30 days (1.50; 1.38,1.64) and remained elevated despite steady decline over 1 year (1.15; 1.10,1.21). In contrast, in hyperlipidemia, the risk of ASCVD hospitalizations was elevated in the first 30 days (2.44; 2.24, 2.65) and steadily increased over the first year (2.78; 2.67, 2.90). The following risk factors were positively associated with ASCVD hospitalizations after COPD hospitalization over the entire 1-year: peripheral vascular disease (1.27), congestive heart failure (1.17), valvular heart disease (1.17), renal disease (1.14), diabetes (1.13), hypertension (1.09) and index year 2018 (1.10) and 2019 (1.08).

**Figure 4:**
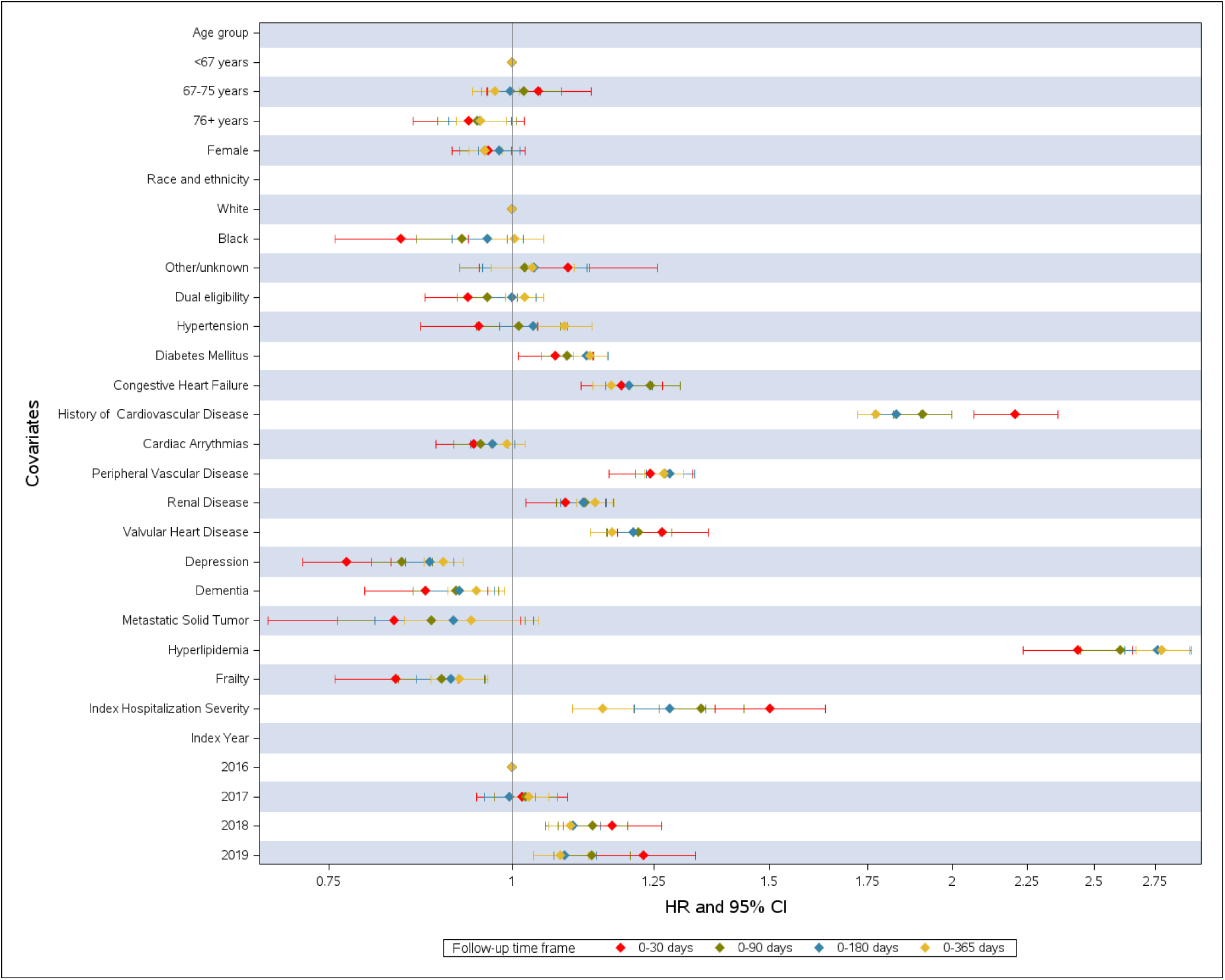
Forest plot showing adjusted hazard ratio estimates for association between CVD composite outcome and predictors of interest over 30, 90, 180, and 365 days from date of discharge; estimates were obtained from separate Cox proportional hazard models for each time frame (0-30 days, 0-90 days, 0-180 days, and 0-365 days; indicated in plot with color key).

## DISCUSSION

In this study, we performed a retrospective analysis using Medicare 100% fee-for-service inpatient claims data to study timing of ASCVD hospitalization risk and risk factors for ASCVD hospitalization among older adults hospitalized for COPD. We observed no increased risk of an ASCVD hospitalization composite outcome in the after-COPD hospitalization period relative to the before-COPD hospitalization period after adjusting for patient characteristics. However, we identified several important interactions, including multiple subgroups (age +76 years, women, and very severe COPD hospitalization) who experienced increased risk of ASCVD hospitalization after-COPD hospitalization relative to before-COPD hospitalization. In addition, we identified 10 risk factors, namely hyperlipidemia and history of cardiovascular disease, which were at highest risk of ASCVD hospitalization after-COPD hospitalization. Collectively, these findings suggest that, while risk of ASCVD hospitalization is not elevated after-COPD hospitalization relative to before-COPD hospitalization after accounting for patient characteristics, there appears to be increased risk of ASCVD hospitalization following COPD hospitalization among high-risk subgroups.

We found the risk of an ASCVD hospitalization composite outcome was not higher post– vs. pre-COPD hospitalization. This finding contrasts with results from numerous studies demonstrating higher risk of ASCVD events after COPD exacerbations^19^. However, there are several key differences between these prior studies and our study which may explain these conflicting results. First, prior studies investigated ASCVD event risk among patients with either a history of ASCVD events^5, 8^, history of cardiovascular disease or ASCVD risk factors^6^. In contrast, our study focused on a broader population, not enriched for cardiovascular disease, to minimize selection bias and enhance generalizability. Additionally, prior studies demonstrated increased risk of ASCVD events within 49 days after COPD exacerbation^5, 8, 20^.

Whereas we examined risk in the 1 year period after COPD hospitalization. Considering the proportional hazard assumption was violated, it is plausible that the longer follow up time period attenuated the signal for ASCVD hospitalization risk in our study. However, while the risk of ASCVD events after-COPD hospitalization has been shown to be the highest immediately following COPD exacerbation, ASCVD event risk has been reported to remain two-fold higher 1-year after COPD exacerbation^6^. Lastly, the primary outcome of a CVD composite event outcome differed by study. A post-hoc analysis of the SUMMIT study included cardiovascular death as an adjudicated event in the composite CVD endpoint which accounted for 39% of all CVD events^6^. We were unable to include cardiovascular death in our composite end point given cause of death data is not available in Medicare files. Therefore, it is possible that we underestimated ASCVD risk after-COPD hospitalization given that we were unable to include cardiovascular death in our composite endpoint. With that being said, cardiovascular death could only be observed in the post-COPD hospitalization period as a patient who experienced CVD death in the pre-COPD hospitalization period would not be included in the study cohort.

Thus, by not including CVD death as an endpoint in our composite outcome, we reduced the potential risk of bias.

Importantly, we identified several key subgroups who had higher ASCVD hospitalization risk in the after-COPD hospitalization period compared to the before, including patients with very severe COPD hospitalizations, women, and age 76+ years. Previous studies have demonstrated the risk of ASCVD events is highest among patients requiring hospitalization compared to all exacerbations^5, 8, 21^, with a reported 6-fold higher risk within the first 30 days after exacerbation^6^. Our finding that very severe COPD hospitalizations (i.e., those requiring invasive mechanical ventilation) were at higher risk of ASCVD events compared to less severe COPD hospitalizations is consistent with previous findings demonstrating a positive correlation between COPD exacerbation severity and subsequent ASCVD hospitalization risk. Our finding that female sex was associated with higher risk of ASCVD hospitalizations after COPD hospitalization is likely, at least in part, explained by ASCVD being under-recognized, underdiagnosed, and undertreated in women^22^. Lastly, our finding that adults age 76+ were associated with higher risk of ASCVD hospitalizations after COPD hospitalization is notable because despite being at highest risk for atherosclerotic cardiovascular disease events, older adults are known to suffer from treatment gaps and less aggressive preventive strategies than younger populations^23, 24^. Thus, the heightened risk observed here represents another opportunity for targeted preventive efforts in the geriatric population. Our findings that invasive mechanical ventilation during COPD hospitalization, female sex, and age 76+ are associated with higher ASCVD hospitalization risk after COPD hospitalization suggests that the exacerbation event may represent a critical opportunity for additional risk stratification or treatment intensification to prevent future ASCVD hospitalizations in these high-risk subgroups.

We found 10 risk factors that were significantly associated with higher risk of ASCVD hospitalization after-COPD hospitalization. Hyperlipidemia (2.78; 2.67, 2.90) and history of cardiovascular disease (1.77; 1.72 1.83) were associated with the greatest risk of ASCVD hospitalizations 1-year after COPD hospitalization. Our findings suggest that patients with these characteristics may be at the highest risk of ASCVD hospitalizations after COPD hospitalization. In this conceptual model, COPD hospitalization serves as a risk promoting event, leading to further increased risk of ASCVD hospitalization among patients whom at baseline are high-risk. The 10 characteristics we identified as risk factors for ASCVD hospitalizations after COPD hospitalization could be used to inform the design of future studies investigating risk stratification and therapeutic interventional trials targeting patient populations at highest risk for ASCVD hospitalization.

Our study has several strengths including use of Medicare 100% LDS database which provided a large, generalizable population for analysis. To identify our cohort we leveraged previous approaches using Medicare data published in high-impact journals^11, 12^ and previously validated and published coding algorithms to characterize clinical events and comorbidities in our cohort^15, 16, 25^. We extended prior work and minimized selection bias by including beneficiaries with and without a prior history of CVD, which supports generalizability of our findings. We limited our primary composite ASCVD outcome to acute, atherosclerotic cardiovascular disease events resulting in hospitalization opposed to other studies which also included heart failure^21^, pulmonary embolism and atrial fibrillation^26^. However, as the Medicare 100% LDS database does not include outpatient or carrier claims, our comorbidity definitions were limited to diagnostic codes from inpatient claims. Additionally, the Medicare dataset does not include medications. Therefore, residual confounding may be present as we could not account for most notably CVD medications. However, we expect that most patients in such a large, generalizable cohort received standard of care treatment^27^. Moreover, there is a lack of information on services not covered under Medicare, as well as Medicare beneficiaries enrolled in Medicare Advantage.

Among a broad population of older adults, we found no increased risk of atherosclerotic cardiovascular disease events leading to hospitalization after-COPD hospitalization relative to before-COPD hospitalization. However, we identified 3 subgroups (age 76+, women, very severe COPD hospitalization) and 10 risk factors associated with increased risk of ASCVD hospitalizations after-COPD hospitalization with hyperlipidemia and history of cardiovascular disease associated with the greatest risk. Our findings suggest that ASCVD hospitalization risk may be increased after COPD hospitalization among select patients with a high-risk clinical phenotype. This characterization of the COPD exacerbation populations at highest ASCVD hospitalization risk can inform the development of novel ASCVD risk assessment and reduction strategies.

## Data Availability

Data will be made available upon formal request and consideration.

## ACKNOWLEDGEMENTS

Study authors wish to thank Melissa A. Greiner, MS for her expertise and contributions towards study design and biostatistical analysis prior to retirement and Chantelle Hardy, MPH for her initial support in the role of project leader.

## SOURCES OF FUNDING DISCOSURES

CLM reports research support from the American Lung Association Early Career Investigator Award and The Lang Family COPD Research Fund to complete this project. CLM receives research funding unrelated to this project from the Patient-Centered Outcomes Research Institute, AstraZeneca, and the National Institute on Aging/National Institutes of Health from R03AG082878-01(GEMSSTAR award).

Consulting fees with COPDFoundation and Wellinks. The American Lung Association and Lang Family Fund were not involved in the research study or in the presentation of our findings. MGN reports current research support from the American College of Cardiology Foundation supported by the George F. and Ann Harris Bellows Foundation, the Patient-Centered Outcomes Research Institute, the Yale Claude D. Pepper Older Americans Independence Center (P30AG021342), and the National Institute on Aging/National Institutes of Health from R03AG074067 (GEMSSTAR award).

Consulting fees from HeartFlow Inc., Merck. WSJ reports research support from Bayer, Boehringer Ingelheim, Merck, National Institutes of Health, and Patient Centered Outcomes Research Institute. ECO receives research funding unrelated to this project from BMS, Pfizer, Patient-Centered Outcomes Research Institute, and Novartis. The remaining authors have no disclosures relevant to this work.

